# The Relationship Between Neighborhood Safety, Paternal Presence and Somatic Symptoms of Dissociation in Children

**DOI:** 10.1101/2024.03.07.24303927

**Authors:** Jenna Pacelli

## Abstract

**Introduction:** Child exposure to prolonged stressful home environments can lead to life-long patterns that are later misdiagnosed in adulthood as anxiety disorders, ADHD and depressive disorders, among others. In theory, paternal involvement and neighborhood safety are strong buffers against toxic stress and therefore dissociative symptoms in children. The relationship between paternal involvement and rates of violence in the neighborhood and rates of dissociative symptoms in children are not yet fully understood. This study investigates the relationship between paternal involvement or neighborhood safety on somatic symptoms of dissociation in children.

**Methods:** This was a retrospective study based on the open-source dataset from Welfare, Children, and Families: A Three-City Study, Dataset 0005 (Angel et al., 2012). A cross-tab analysis investigated the relationship between 1) the likelihood of neighborhood assaults and 2) how often the child saw/spoke to their father in the last year with the likelihood of a child having a) concentration issues, b) confusion, c) being lost in thought, d) hearing voices and e) staring blankly.

**Results:** Results show that children who lived in neighborhoods where there were more muggings and assaults were more likely to be staring blankly and have concentration problems. Children who saw or talked to their father less were more likely to have concentration problems. Lastly, children who saw their fathers less often were more likely to hear voices that were not there.

**Conclusion:** This study showed a relationship between rates of violence in the neighborhood and paternal presence and symptoms of dissociation in children. This suggests that the lived experience of the child affects their cognition and can be impacted by living in unsafe areas and not being able to see or speak with their father. Dissociative symptoms are linked with an increased risk of substance abuse, suicide attempts, heart disease, cancer, and skeletal fractures. Understanding factors that affect dissociation will provide opportunities to prevent childhood dissociation.

## Introduction

Neuroception is the subconscious system in the brain, body and nervous system that allows mammals to detect threat and safety in their environment (Porges, 2011, pp. 11-18). Ideally, a child’s environment allows them to feel a sense of safety. If not, the child will take defensive measures that affect all her body’s systems. Faulty neuroception - the ability to accurately perceive whether or not the environment is safe - might lie at the root of many psychiatric disorders (Porges, 2011, pp. 11-18). For example, areas in the temporal cortex that may inhibit fight, flight, or freeze reactions are not active in people with autism and schizophrenia. Individuals with anxiety and depression have difficulties in regulating heart rate and exhibit lower heart rate variability (HRV), (Porges and Furman, 2011, pp. 17). Abused and institutionalized children with reactive attachment disorder tend to be over or under-inhibited - signs that their sympathetic and parasympathetic nervous systems are overly reactive to their environment, despite potential new signs of safety (Porges and Furman, 2011, pp. 17).

Dissociation is a clinical psychiatric symptom defined as the fragmentation and splitting of the mind and perception of the self and body (Scaer, 2001). It presents clinically as altered perceptions and behavior through derealization (the feeling of the surrounding environment not being real), depersonalization (the feeling of the person not being real), distortions of time, space and body, and is linked to freeze or immobility states in the nervous system.

This study investigates the relationship of paternal presence and neighborhood safety with somatic symptoms of dissociation in children. The results will provide knowledge on the effects of where a child lives, how often they spend time with their father on their cognition and development and potential for developing mental health disorders later in life. This study reviews the literature on paternal involvement, adverse childhood experiences, the neurophysiology of trauma and dissociation in children and the economic costs of these issues on society. In turn, we can use this knowledge to recognize somatic symptoms in children, explore variables that may lower dissociative symptoms and improve the economic burden of mental health disorders on nations globally.

### 1.2 Somatic Symptoms of Dissociation in Children

Young children show signs of dissociation in somewhat distinct ways from adults. Children have limited motor and expressive language skills (Levine, 2019, pp. 43). They cannot verbally recount what is happening or has happened to them. They will show trauma symptoms through play, sleep patterns, bed wetting, staring into space vacantly, altered activity levels, outsized emotional reactions and somatic complaints such as headaches (Levine, 2019, pp. 43). In this study, I analyzed the following somatic signs of dissociation as independent variables: struggles with concentration and confusion, feeling lost in a fog, daydreaming or being lost in thought, hearing voices that are not there or relying heavily on “imaginary friends,” which can be an early sign of Dissociative Identity Disorder (DID), thought to be the most severe form of PTSD. Patients with PTSD have also been shown to have deficits in short-term memory (Bremner et al., 1993). Children with complex trauma (cPTSD) score higher than children with no trauma or single-event trauma on all scales of anxiety, depression, anger, PTSD, immaturity, health concerns, familial alienation and psychoticism (Luoni et al., 2018).

Infants are the most prone to dissociation in the face of stress as they do not yet have access to fight or flight responses in their bodies. The infant will detach, withdraw, fuss, cry, protest, have vacant looks in their eyes, and flat affect or facial expressions (Levine, 2019, pp. 44). Unresolved hyperarousal in the nervous system will lead to a dorsal vagal shutdown, dissociation, freeze or hypoarousal in the nervous system. This causes the tissues of the body to constrict, tense, and shut down (Levine, 2019, pp. 46). Left unattended, these chronic freeze states can lead to further core dysregulation of the nervous system and health issues that come from the body being in a chronic state of toxic stress.

### 1.3 Adverse Childhood Experiences

It has been shown that fostering strong, responsive relationships between children and their caregivers is a strong buffer for children from the effects of toxic stress in their environment. All these conditions create toxic stress in the body of the child. Stress responses are meant to be time-limited evolutionarily and then complete once the stressful event is over. Violence in the neighborhood is a strong indicator of the safety of their overall environment. The ACEs are physical abuse, sexual abuse, emotional neglect and abuse, physical neglect, having a mentally ill, depressed or suicidal person in the home, having a drug addicted or alcoholic family member in the home and witnessing domestic violence against the mother (Felitti et al., 1998).

### 1.4 Paternal Involvement

Infants of highly involved fathers, as measured by the amount of interaction, higher levels of play and caregiving activities are more cognitively competent at six months and achieve higher scores on the Bayley Scales of Infant Development (Allen, Daly, 2007). The effects are distinct from the involvement of mothers: fathers’ talk with toddlers is characterized by more “wh-” (what, where, why) questions, which invites the child to talk more, use more diverse vocabulary and speak in longer phrases (Allen & Daly, 2007). School-aged children of more involved fathers perform better in school and have greater success overall throughout the course of their lives (Allen & Daly, 2007).

Yet much is still not fully understood about the relationship between neighborhood safety and childhood somatic symptoms of dissociation, and paternal presence and childhood somatic symptoms of dissociation. The current study investigates these relationships in children using neighborhood mugging and assault likelihood to measure neighborhood safety, how often a child saw or spoke to their father in the last year as a measure of paternal presence and having issues with concentration, confusion, daydreaming, hearing voices and blank stares as measures of somatic symptoms of dissociation. I hypothesize that there will be a significant relationship between neighborhood safety and somatic symptoms of dissociation, and between paternal presence and somatic symptoms of dissociation.

## 2. Methods

### 2.1 Participants

The sample size of this study included 1835 children between the ages of 0 and 20 (*M* = 12.17). Within our sample 50.5% (N= 928 were male), and 49.45% (N= 907) were female. The data collection was conducted amongst low-income families in Boston, San Antonio and Chicago. There were three waves of data collection spanning March 1999 and May 2006 where researchers interviewed caregivers and older children and assessed younger children. Forty percent of the families were receiving cash welfare payments at the time of the interviews and had incomes less than 200 percent of the government poverty line. All interviews were conducted in person, using computer-assisted personal interviews, computer-assisted phone interviews, face-to-face interviews and phone interviews.

### 2.2 Exclusion Criteria

Subjects were excluded from a measure when there was missing data for that variable.

### 2.3 Study Design

This was a retrospective study based on the open-source dataset from Welfare, Children, and Families: A Three-City Study, Dataset 0005 (Angel et al., 2012). The project examined the ways in which families coped with welfare reform. The primary focus of the study was to determine how those dynamics affected the lives of children, with an emphasis on their health and wellness, social-emotional development and subsequent need of social services.

### 2.4 Data Variables

The current study investigates the relationship between variables related to safety and relational attachment inside and outside the home and symptoms of somatic dissociation of the child. There were three independent variables. The first looked at the safety of the neighborhood by looking at the likelihood of muggings and assaults in their neighborhood (not a problem, somewhat of a problem, a big problem). The second independent variable measured paternal presence in the child’s life as indicated by how often the child talked to their father in the last 12 months (never in the past 12 months, every few months, once a month or more, almost every day, never seen their father and father is in the household). The third independent variable measured how often the child saw their father in the last 12 months (never seen the father, never in the past 12 months, once a month or more, once a week or more, almost every day, father in the household.

The five dependent variables measured somatic symptoms of dissociation in the child. Specifically, the dependent variables were the likelihood of a child 1) having issues with concentration, 2) having issues with confusion or being in a fog, 3) daydreaming or being lost in thought, 4) hearing voices that aren’t there and 5) staring blankly. All the dependent variables were measured as not true, somewhat or sometimes true, and often true or very true.

### 2.5 Statistical Analysis

To determine whether a difference between observed and expected data was due to a relationship between our independent and dependent variables or chance, a chi-squared test of independence was performed. I performed Rao-Scott adjustments due to complex survey design. Significance was set at p < 0.05 . With three independent variables and five dependent variables, a total of 15 statistical tests were performed.

## 3. Results

I examined the relationship of three independent variables: 1) the likelihood of neighborhood muggings, 2) How often the child talked to their father in the last 12 months and 3) How often the father saw the child in the last 12 months and five dependent variables: the likelihood of a child having issues with 1) concentration, 2) having issues with confusion or being in a fog, 3) daydreaming or being lost in thought, 4) hearing voices that aren’t there and 5) staring blankly.

### 3.1 The relationship between the likelihood of neighborhood muggings and somatic symptoms of child dissociation

A chi-squared test of independence was performed to examine the relationship between if neighborhood assaults and muggings were a problem (not a problem, somewhat of a problem, a big problem) and if a child had problems concentrating (not true, somewhat/sometimes true, very/often true). The relationship between these variables was significant, *X^2^* (4, N= 1,835) =9.98, Rao-Scott Adjusted Pearson’s F =2.49, *p* = .04 (Fig. 1). Children who lived in neighborhoods where muggings were a frequent problem were more likely to often struggle with concentration, whereas children who lived in neighborhoods where muggings were not a problem did not struggle with concentration.

A chi-squared test of independence was performed to examine the relationship between if neighborhood assaults and muggings were a problem (not a problem, somewhat of a problem, a big problem) and if a child was observed as being confused or in a fog (not true, sometimes true, very true or often true). The relationship between these variables was not significant, *X^2^* (4, N= 1,835) = 4.8, Rao-Scott Adjusted Pearson’s F =1.20, *p* = .31 (Fig. 2). These results suggest that there was no significant relationship between if neighborhood assaults and muggings were a problem and if a child had problems with being confused or in a fog.

A chi-squared test of independence was performed to examine the relationship between if neighborhood assaults and muggings were a problem (not a problem, somewhat of a problem, a big problem) and if a child was observed as daydreaming or being lost in thoughts (not true, somewhat or sometimes true, very true or often true). The relationship between these variables was not significant, *X^2^* (4, N= 1,835) = 5.52, Rao-Scott Adjusted Pearson’s F = 1.38, p = 0.24 9 (Fig. 3). This suggests that there was no significant relationship between if neighborhood assaults and muggings were a problem and if a child was observed daydreaming or being lost in thoughts.

A chi-squared test of independence was performed to examine the relationship between if neighborhood assaults and muggings were a problem (not a problem, somewhat of a problem, a big problem) and if a child hears voices that are not there (not true, somewhat or sometimes true, very true or often true). The relationship between these variables was not significant *X^2^*(4, N= 1,835) = 6.48, Rao-Scott Adjusted Pearson’s F = 1.62, p = 0.17 (Fig. 4) This suggests that there was no significant relationship between if neighborhood assaults and muggings were a problem and if a child hears voices that are not there.

A chi-squared test of independence was performed to examine the relationship between if neighborhood assaults and muggings were a problem (not a problem, somewhat of a problem, a big problem) and if a child was observed staring blankly (not true, somewhat or sometimes true, very true or often true). The relationship between these variables was significant *X^2^* (4, N= 1,835) = 14.41, Rao-Scott Adjusted Pearson’s F = 3.60, p = 0.01 (Fig. 5). Children who lived in neighborhoods where assaults and muggings were a problem were more likely to be observed staring blankly.

### 3.2 The relationship between how often the father talked to the focal child in the last 12 months and somatic symptoms of child dissociation

A chi-squared test of independence was performed to examine the relationship between how often the father talked to the focal child in the last 12 months (never in the past 12 months, every few months, once a month or more, almost every day, never seen their father and father in the household) and if the child struggled with concentration (not true, somewhat/sometimes true, very/often true). The relationship between the variables was significant *X^2^* (12, N= 1,835) = 22.07, Rao-Scott Adjusted Pearson’s F 1.84, p = 0.04 (Fig. 6). Children who had never seen their father were more likely to struggle to concentrate. Children whose fathers were in the household were less likely to struggle with concentration.

A chi-squared test of independence was performed to examine the relationship between how often the father talked to the focal child in the last 12 months (never in the past 12 months, every few months, once a month or more, almost every day, never seen their father and father in the household) and if the child was observed as being confused or in a fog (not true, sometimes true, very true or often true). The relationship between these variables was not significant *X^2^* (12, N= 1,835) = 13.37, Rao-Scott Adjusted Pearson’s F = 1.11, p = 0.34 (Fig. 7). This suggests that there was no significant relationship between how often the father talked to the focal child and if the child was observed as being confused or in a fog.

A chi-squared test of independence was performed to examine the relationship between how often the father talked to the focal child in the last 12 months (never in the past 12 months, every few months, once a month or more, almost every day, never seen their father and father in the household) and if the child daydreams or is observed as lost in thought (not true, somewhat or sometimes true, very true or often true). The relationship between these variables was not significant *X^2^* (12, N= 1,835) = 17.34, Rao-Scott Adjusted Pearson’s F = 1.45, p = 0.14 (Fig. 8). This suggests that there was no significant relationship between how often the father talked to the focal child and how often the child was seen daydreaming or lost in thought.

A chi-squared test of independence was performed to examine the relationship between how often the father talked to the focal child in the last 12 months (never in the past 12 months, every few months, once a month or more, almost every day, never seen their father and father in the household) and if the child hears voices that are not there (not true, somewhat or sometimes true, very true or often true). The relationship between these variables was non-significant but trending *X^2^* (12, N= 1,835) = 13.81,

Rao-Scott Adjusted Pearson’s F = 1.73, p = 0.05 (Fig. 9). This suggests that while there was no significant relationship between how often the father talked to the focal child and if the child heard voices that were not there, there was an emerging trend that indicates some relationship.

A chi-squared test of independence was performed to examine the relationship between how often the father talked to the focal child in the last 12 months (never in the past 12 months, every few months, once a month or more, almost every day, never seen their father and father in the household) and if the child was observed staring blankly (not true, somewhat or sometimes true, very true or often true). The relationship between these variables was not significant *X^2^* (12, N= 1,835) = 15.14, Rao-Scott Adjusted Pearson’s F = 1.51, p = 0.11 (Fig. 10). This suggests that there was no significant relationship between how often the father talked to the focal child and if the child was observed staring blankly.

### 3.3 The relationship between how often the father has seen the focal child in the last 12 months and somatic symptoms of dissociation

A chi-squared test of independence was performed to examine the relationship between how often the father has seen the focal child in the last 12 months (never seen the father, never in the past 12 months, once a month or more, once a week or more, almost every day, father in the household) and if the child struggles with concentration (not true, somewhat/sometimes true, very/often true). The relationship between these variables was significant *X^2^* (12, N= 1,835) = 22.17, Rao-Scott Adjusted Pearson’s F = 1.85, p = 0.04 (Fig. 11). This suggests that children who saw their father more often in the past 12 months were able to concentrate better and those who never see their father struggled the most with concentration.

A chi-squared test of independence was performed to examine the relationship between how often the father has seen the focal child in the last 12 months (never seen the father, never in the past 12 months, once a month or more, once a week or more, almost every day, father in the household) and if the child was observed being in a fog or confused (not true, sometimes true, very true or often true). The relationship between these variables was not significant, *X^2^* (12, N= 1,835) = 14.41, Rao-Scott Adjusted Pearson’s F = 1.20, p = 0.28 (Fig. 12). This suggests that there was no significant relationship between how often the father sees the focal child and the child being observed in a fog or confused.

A chi-squared test of independence was performed to examine the relationship between how often the father has seen the focal child in the last 12 months (never seen the father, never in the past 12 months, once a month or more, once a week or more, almost every day, father in the household) and if the child was observed daydreaming or lost in thought (not true, somewhat or sometimes true, very true or often true). The relationship between these variables was non-significant but trending *X^2^* (12, N= 1,835) = 21.01, Rao-Scott Adjusted Pearson’s F = 1.75, p = 0.05 (Fig. 13). This suggests that while there was no significant relationship between how often the father has seen his child and if the child is lost in thought, there was an emerging trend that indicates some relationship.

A chi-squared test of independence was performed to examine the relationship between how often the father has seen the focal child in the last 12 months (never seen the father, never in the past 12 months, once a month or more, once a week or more, almost every day, father in the household) and if the child hears voices that are not there. The relationship between these variables was significant *X^2^* (12, N= 1,835) = 15.11, Rao-Scott Adjusted Pearson’s F = 1.89, p = 0.03 (Fig. 14). This suggests that children who saw their father more often were less likely to hear voices that weren’t there and children who saw their father more often were less likely to hear voices that weren’t there.

A chi-squared test of independence was performed to examine the relationship between how often the father has seen the focal child in the last 12 months (never seen the father, never in the past 12 months, once a month or more, once a week or more, almost every day, father in the household) and if the child is observed staring blankly (not true, somewhat or sometimes true, very true or often true). The relationship between these variables was non-significant but trending *X^2^* (12, N= 1,835) = 17.49, Rao-Scott Adjusted Pearson’s F = 1.59, p = 0.09 (Fig. 15). This suggests that while there was no significant relationship between how often the father has seen the child in the last 12 months and if the child was observed staring blankly, there was an emerging trend that indicates some relationship.

## 4. Discussion

The current study investigated the relationship between neighborhood safety and childhood dissociation, and paternal presence and childhood dissociation.

Emotional and physical neglect are two experiences that fall under the adverse childhood experiences. Violence in the neighborhood is a strong indicator of the safety of the overall environment. High childhood neglect and high levels of neighborhood violence are correlated to abnormally high levels of physical stress symptoms in children (Lynch, 1998).

The results of this study showed that children living in neighborhoods with higher incidences of muggings and assault were more likely to struggle with concentration and also to be observed staring blankly. Emerging research has linked neighborhood violence and socio-emotional and threat processing in the brain (Suarez et al., 2024). Exposure to community violence was related to increased amygdala activity. Parenting behavior also modulated these effects in that high parental nurturance buffered the effect of exposure to community violence on amygdala reactivity (Suarez et al., 2024).

Results of this study also show that children who spoke to their father more in the last 12 months were less likely to struggle with concentration. These results were also observed when children saw their father more in the last year. By examining levels of paternal involvement with the focal child as well as the levels of violence in the neighborhood of the child, I am attempting to elucidate the effects of both of these patterns on childhood rates of dissociation. The links between these factors are important because previous research has shown the connection between neighborhood disadvantage and brain function in youth (Marshall et al., 2018). Since parental nurturance is of critical importance on the nervous system health of children, this study also suggests the importance of supporting parents in their ability to parent their children, preventing future generations of trauma.

The results also showed that children who saw their father more in the last year were less likely to hear voices that were not there. Previous research has indicated the strong link between adverse childhood experiences and hallucinations: compared with people with zero ACEs, those with seven or more ACEs had a fivefold increase in the risk of reporting hallucinations (Whitfield et al., 2005). Furthermore, the frequency of ACEs was almost double among schizophrenic patients than in control subjects and a history of multiple ACEs is associated with more severe symptomatology, including increased risk of suicidality (Prokopez et al., 2020).

One finding that was not significant (p = 0.05) but trending toward significance, indicating a relationship was that when children saw their father more often in the last year they were less likely to be observed daydreaming or lost in thoughts. Being lost in thought is a common sign of dissociation and autonomic nervous system (ANS) dysregulation but is often misdiagnosed as ADHD (Levine, 2019, pp. 34). With the skyrocketing prevalence of ADHD diagnoses and therefore prescription of psychotropic stimulants in children at present in the United States, it is of critical importance to examine the underlying nervous system mechanisms of those symptoms. Because parental nurturance and in particular paternal involvement are such strong buffers against the effects of ACEs, it comports with observations that increased paternal involvement would buffer a child from common symptoms of dissociation - having a parent involved in the child’s life allows for life to feel more tolerable, reducing the need for dissociation because the child’s brain needs safe social engagement from their caregivers for proper neural development (Siegel, 2015, pp.34).

Some findings in this study were not statistically significant but were trending near significance. Children who talked to their father less were more likely to be confused, in a fog, daydreaming or lost in thought, hearing voices that weren’t there and staring blankly. Similarly, children who saw their father less often were more likely to be confused, in a fog, daydreaming, lost in thought and staring blankly. Parental nurturance and paternal involvement are clear buffers for these somatic symptoms of trauma. Supporting parents in their own trauma healing and nervous system regulation will therefore support their children to be more regulated, even in the face of violence in their environment.

The limitations of this study and the data therein are that there was missing data for variables in the dataset. A second limitation was that some of the variables including being confused, in a fog, being lost in thought and hearing voices that are not there are more difficult to assess by an outside observer. An experienced clinician trained in the somatic markers of dissociation would be better equipped to observe these symptoms but surveying general lay people such as parents or caregivers or clinicians untrained in somatic dissociation is an inherently flawed method for identifying these internal symptoms of dissociation. Dissociation symptoms are often misunderstood even by clinicians without specialized training in somatic dissociation. Another possible limitation is that children might not report hearing voices that aren’t there because in the case of Dissociative Identity Disorder (DID), children with these symptoms tend to know that it is an abnormal symptom to experience so self-report measures will inherently be inaccurate. Another limitation is that dissociative symptoms are difficult for the person experiencing them to notice and report as well as it is the very nature and function of dissociation to cause the person to be less aware of themselves, their internal experience and their surroundings.

Overall, the results of this study showed a relationship between neighborhood safety and children having concentration problems and staring blankly and between paternal presence and concentration problems, hearing voices and daydreaming.

These results indicate that the lived experience of the child affects their cognition and can be impacted by living in unsafe areas and not being able to see or speak with their father. ACEs are linked with a 4-12 fold increased risk of alcoholism, drug abuse, depression, suicide attempts, a 2-4 fold increase in smoking, poor health, 50 or more sexual partners and sexually transmitted disease and a 1.4 to 1.6-fold increase in physical inactivity and severe obesity (Felitti et al., 1998). A number of these ACEs show a graded relationship to the presence of adult diseases including ischemic heart disease, cancer, chronic lung disease, skeletal fractures and liver disease. There is a strong link between ACEs and many of the leading causes of death (Felitti et al., 1998).

Prolonged exposure to toxic stress leads to a numbing and shutdown of the nervous system. These issues eventually lead to lifelong patterns that are later misdiagnosed in adulthood as anxiety disorder, ADHD, conduct disorders, obsessive compulsive disorder, dissociative disorders, substance abuse disorders and depression amongst many others. The cost of untreated mental illness in the US state of Indiana alone was $4.2 billion annually (Taylor et al., 2023). The estimated economic burden of childhood neglect and abuse in the United States is $428 billion (Peterson et al., 2018). Prevention of mental health disorders and toxic stress caused by Adverse Childhood Experiences consisting of child physical abuse, child sexual abuse, child emotional abuse, emotional neglect, physical neglect, having a mentally ill, depressed, or suicidal person in the home, having a drug addicted or alcoholic family member and witnessing domestic violence against the mother (Felitti et al., 1998) and therefore the burden placed upon the body and nervous system is an issue of global concern (Peterson et al., 2018).

## Data Availability

All data produced are available online at https://www.icpsr.umich.edu/web/DSDR/studies/4701/summary

## Appendix

**Figure 1:**
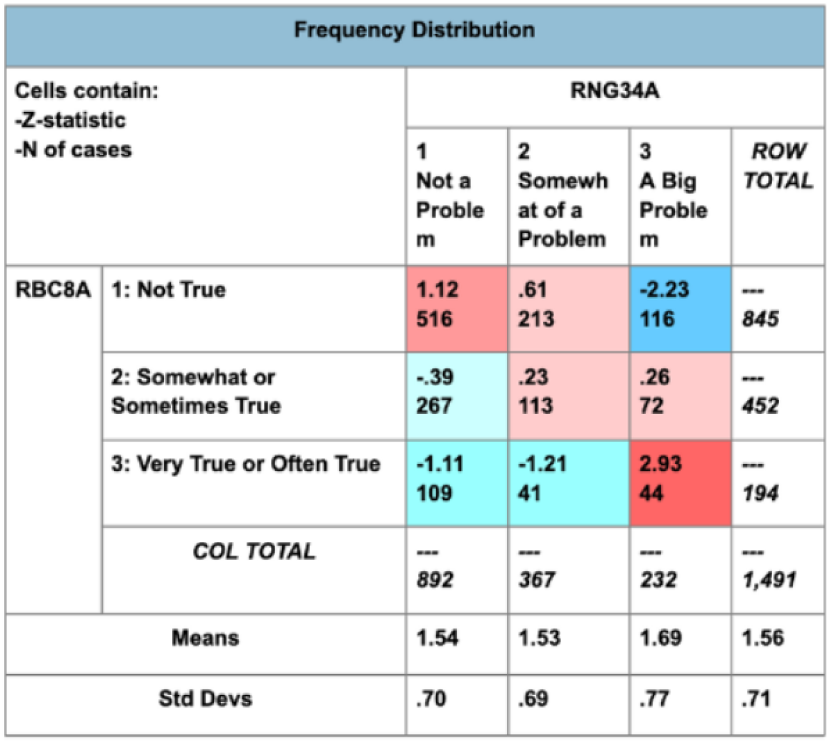
Frequency distribution of chiLd can’t concentrate (RBC8A) and neighborhood assaults/muggings (RNG34A)

**Figure 2:**
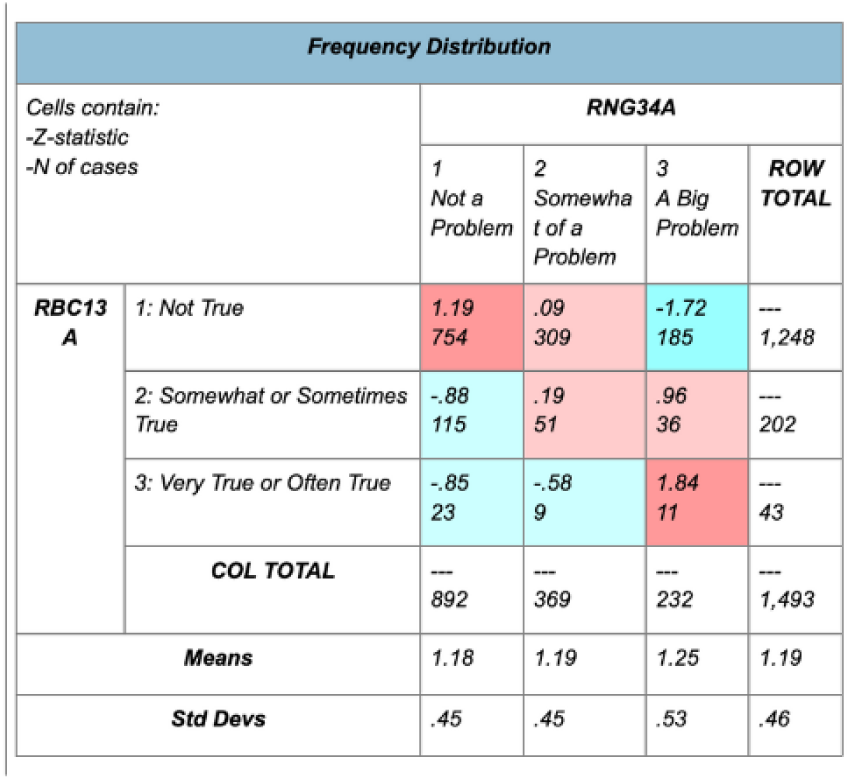
Frequency Distribution of child confused/in a fog (RBC13A) and neighborhood assaults/muggings (RNG34A)

**Figure 3:**
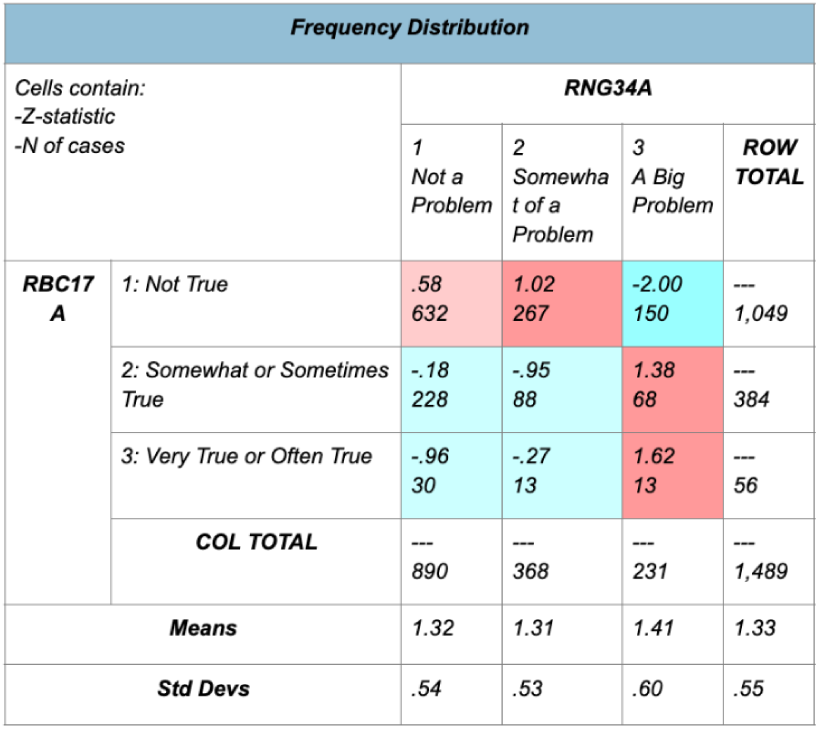
Frequency Distribution of child daydreams/lost in thought (RBC17A) and neighborhood assaults/muggings (RNG34A)

**Figure 4:**
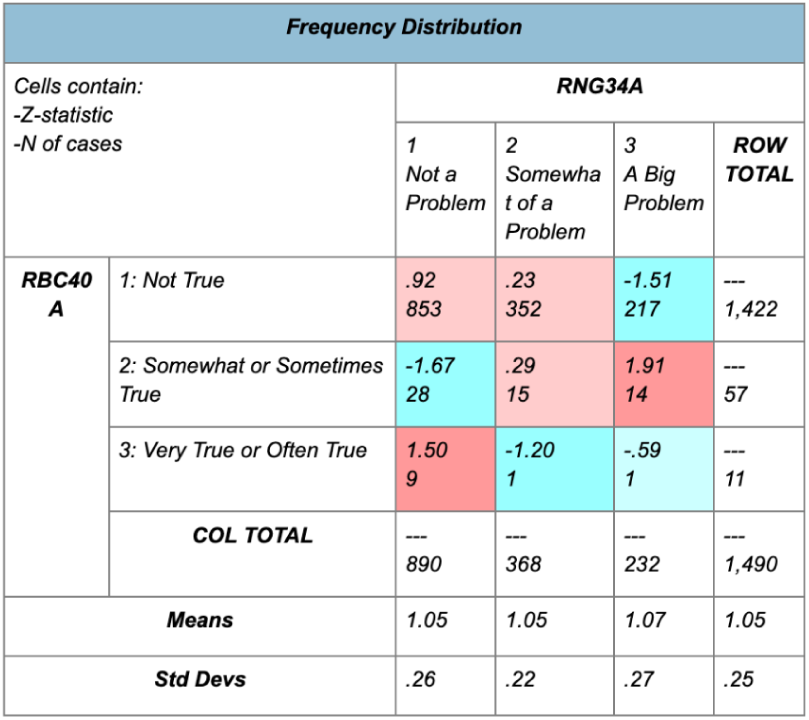
Frequency distribution of child hears voice that aren’t there (RBC40A) and neighborhood assaults/muggings (RNG34A)

**Figure 5:**
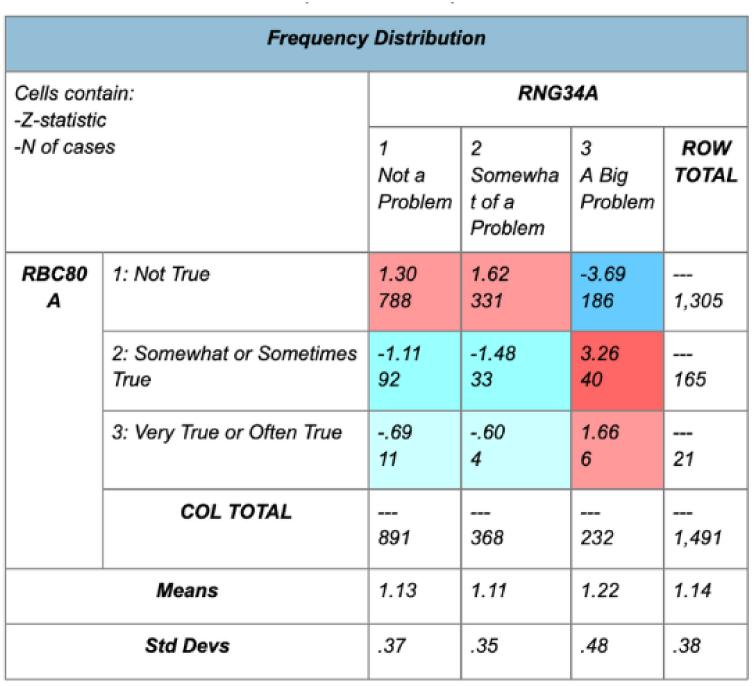
Frequency distribution of child stares blankly (RBC80A) and neighborhood assaults/muggings (RNG34A)

**Figure 6:**
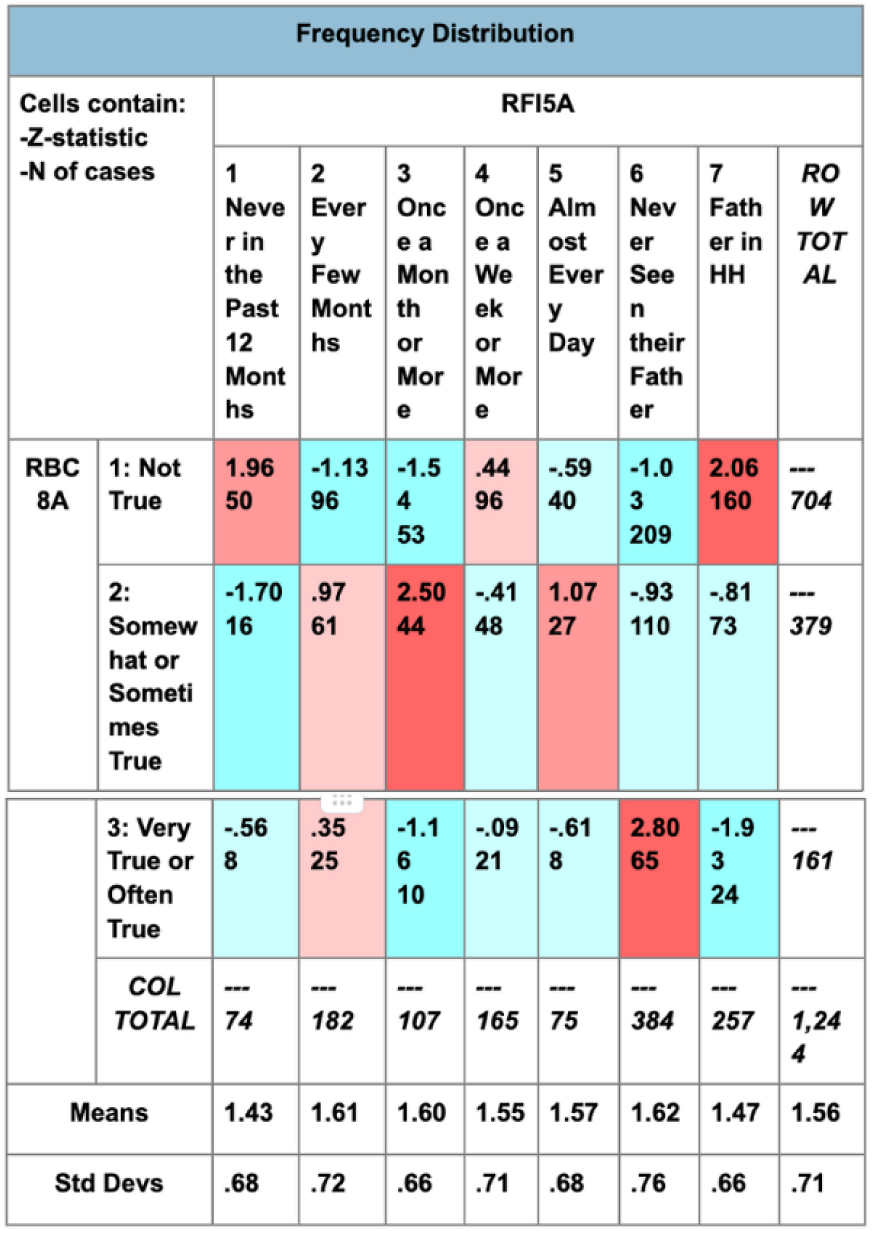
Frequency distribution of child can’t concentrate (RBC8A) and how often the father has talked to the focal child in the last 12 months (RFI5A)

**Figure 7:**
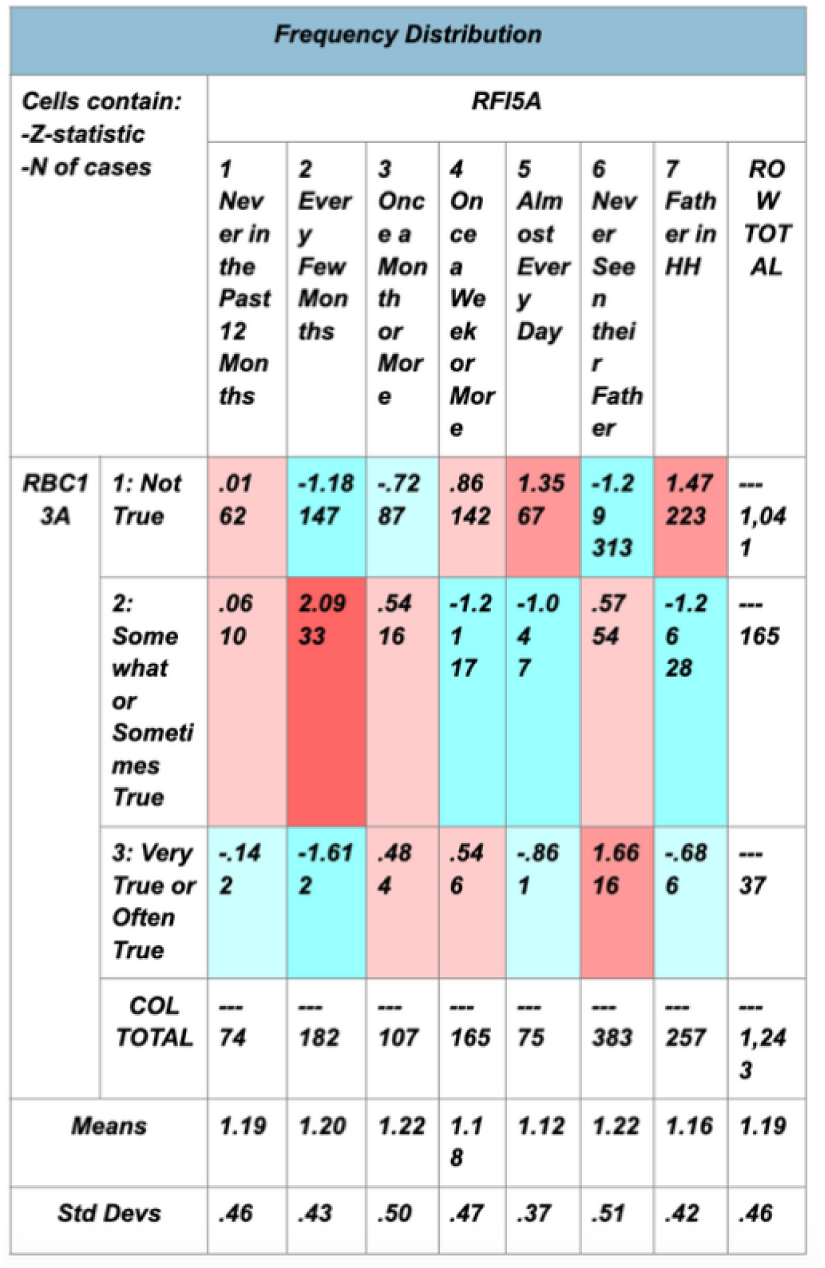
Frequency distribution of child is confused/in a fog (RBC13A) and how often the father has talked to the focal child in the last 12 months (RFI5A)

**Figure 8:**
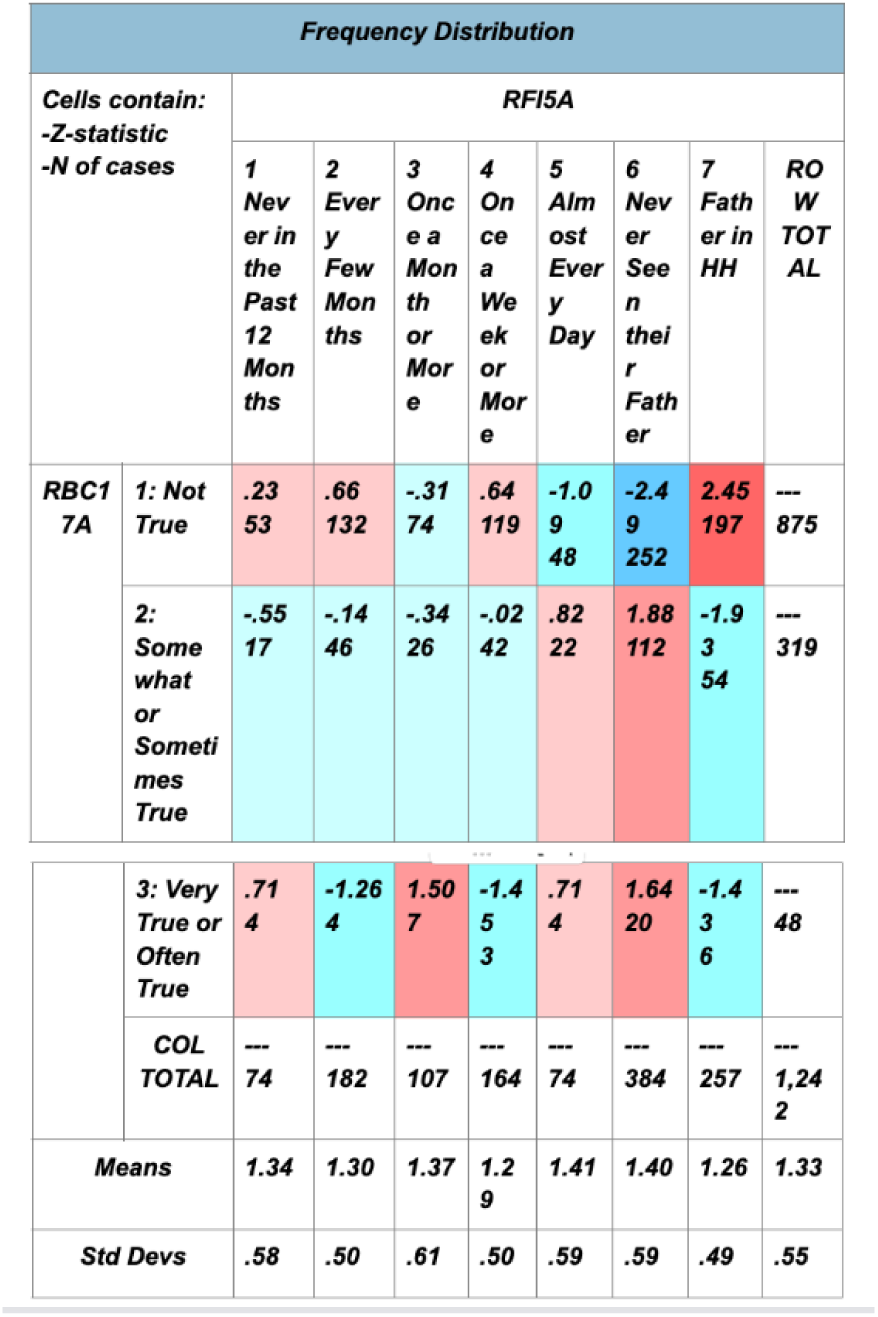
Frequency distribution of child daydreams/is Lost in though (RBC17A) and how often the father has talked to the focal child in the last 12 months (RFISA)

**Figure 9:**
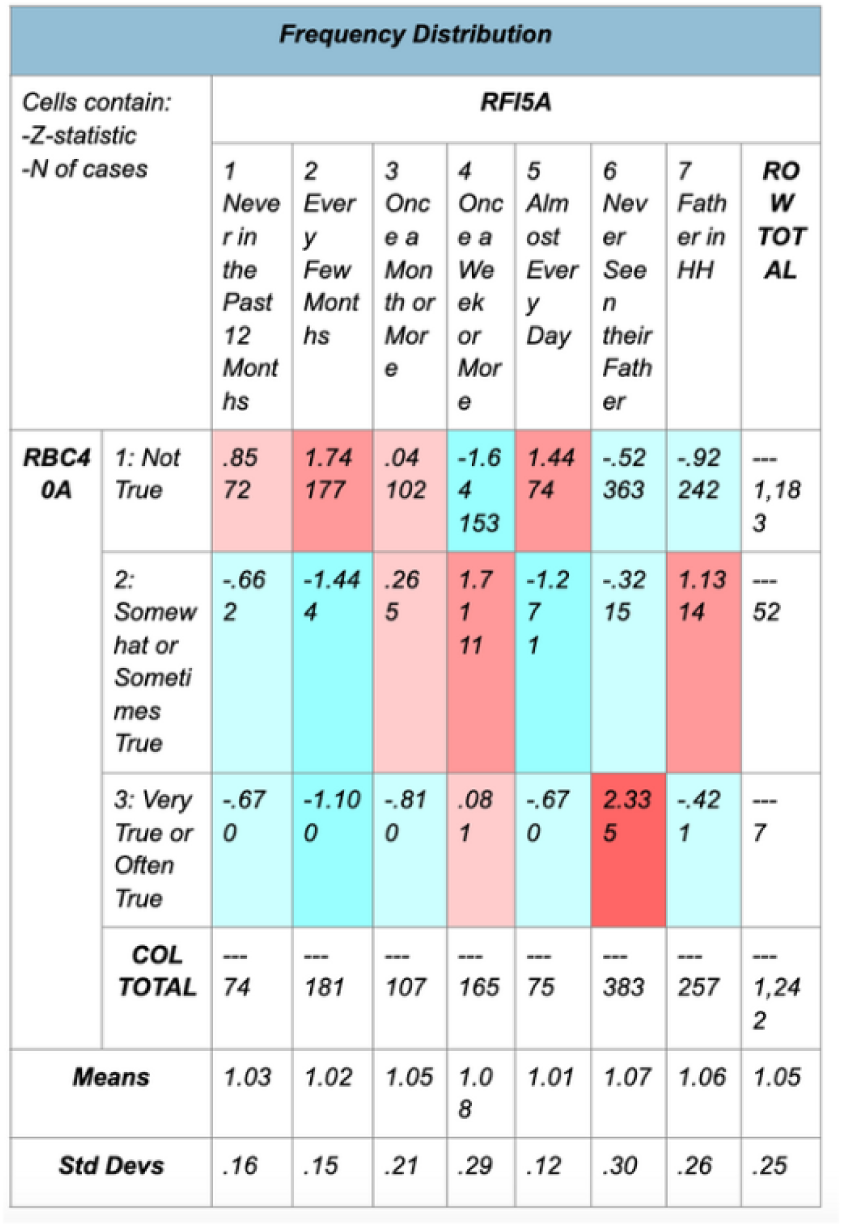
Frequency distribution of child hears voices that aren’t there (RBC40A) and how often the father has talked to the focal child in the last 12 months (RFISA)

**Figure 10:**
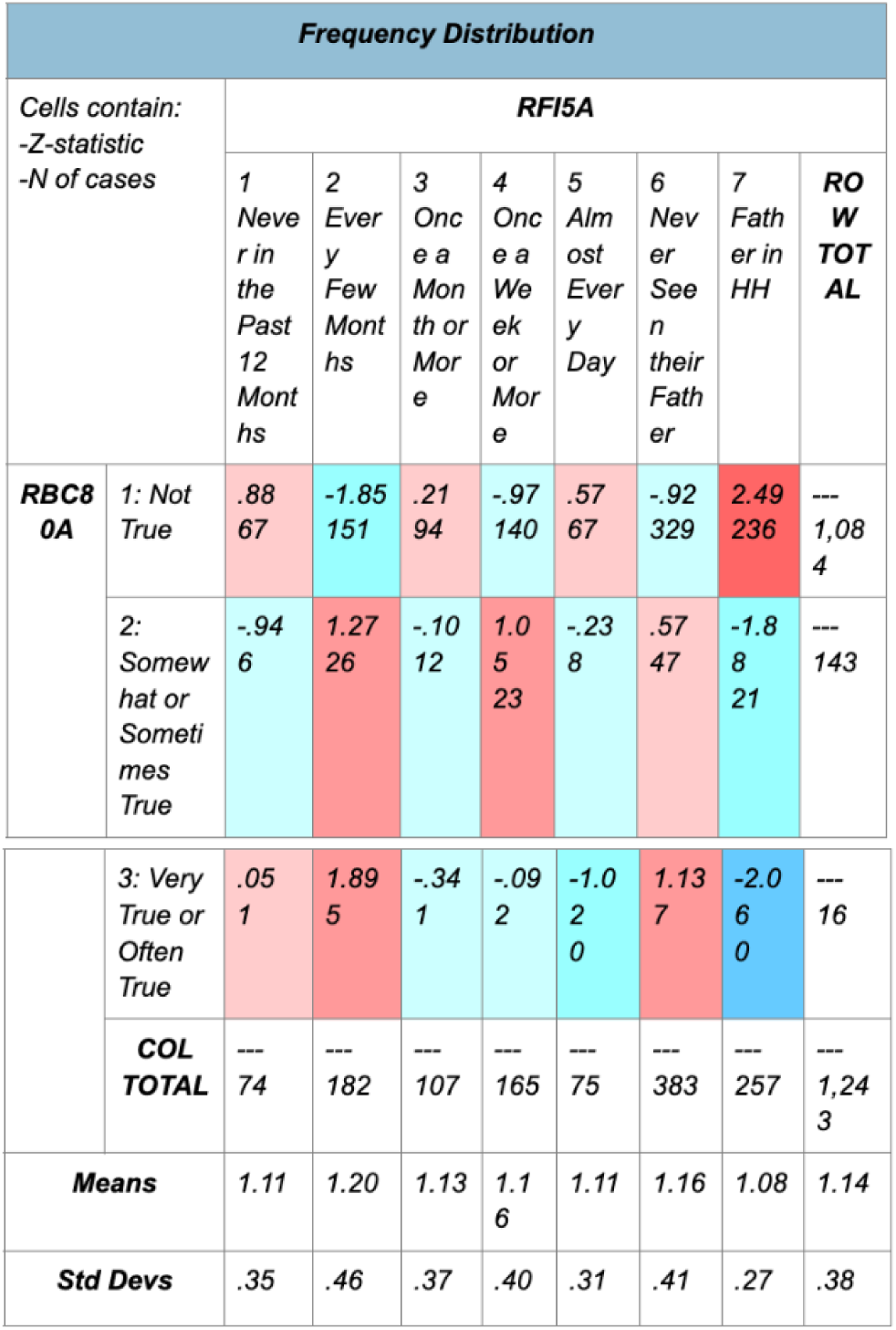
Frequency distribution of child stares blankly (RBC80A) and how often the father has talked to the focal chiLd in the last 12 months (RFI5A)

**Figure 11:**
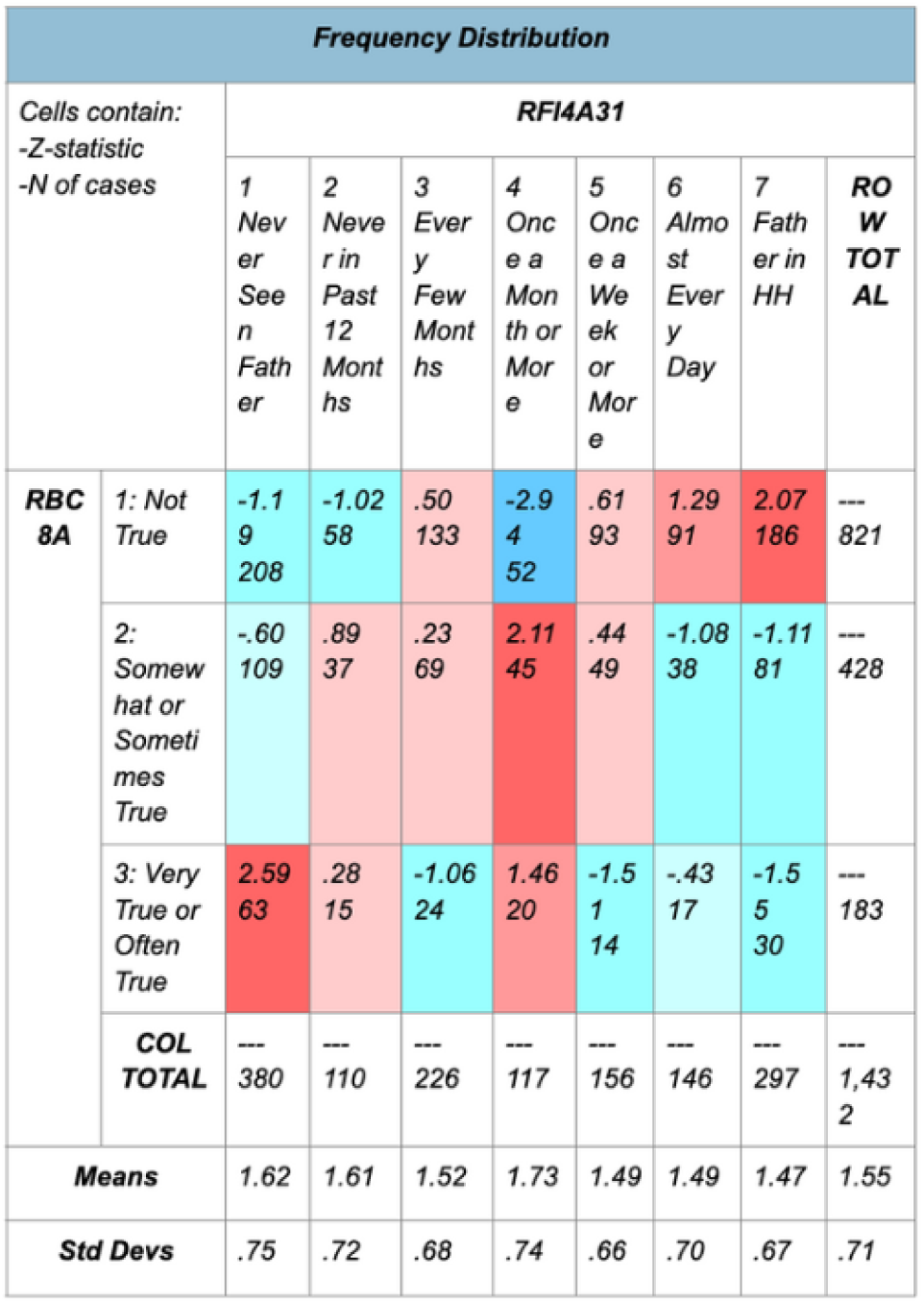
Frequency distribution of child can’t concentrate IRBC8A) and how often the father has seen the focal child in the last 12 months (RFI4A31)

**Figure 12:**
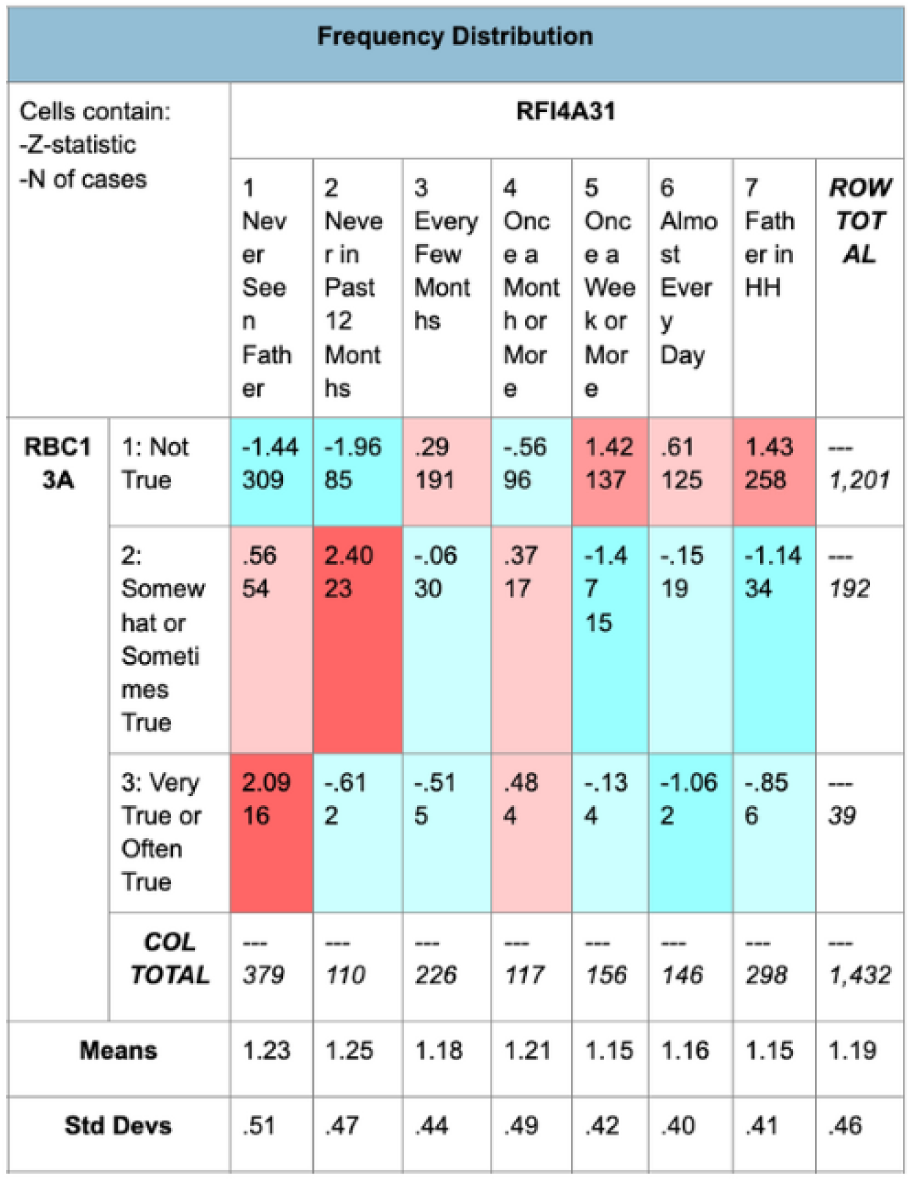
Frequency distribution of child confused/in a fog (RBC13A) and how often the father has seen the focal child in the last 12 months (RFI4A31)

**Figure 13:**
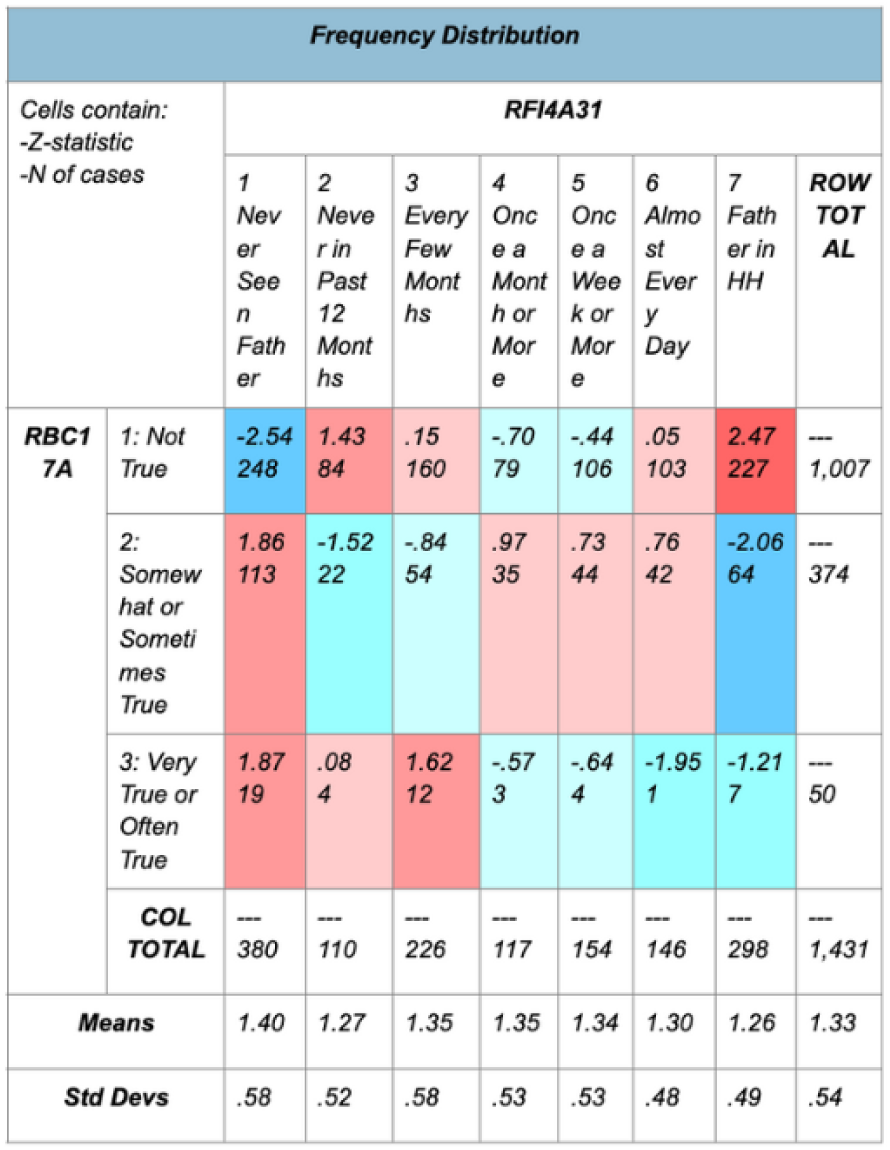
Frequency distribution of child daydreams/is lost in thought (RBC17A) and how often the father has seen the focal child in the last 12 months (RFI4A31)

**Figure 14:**
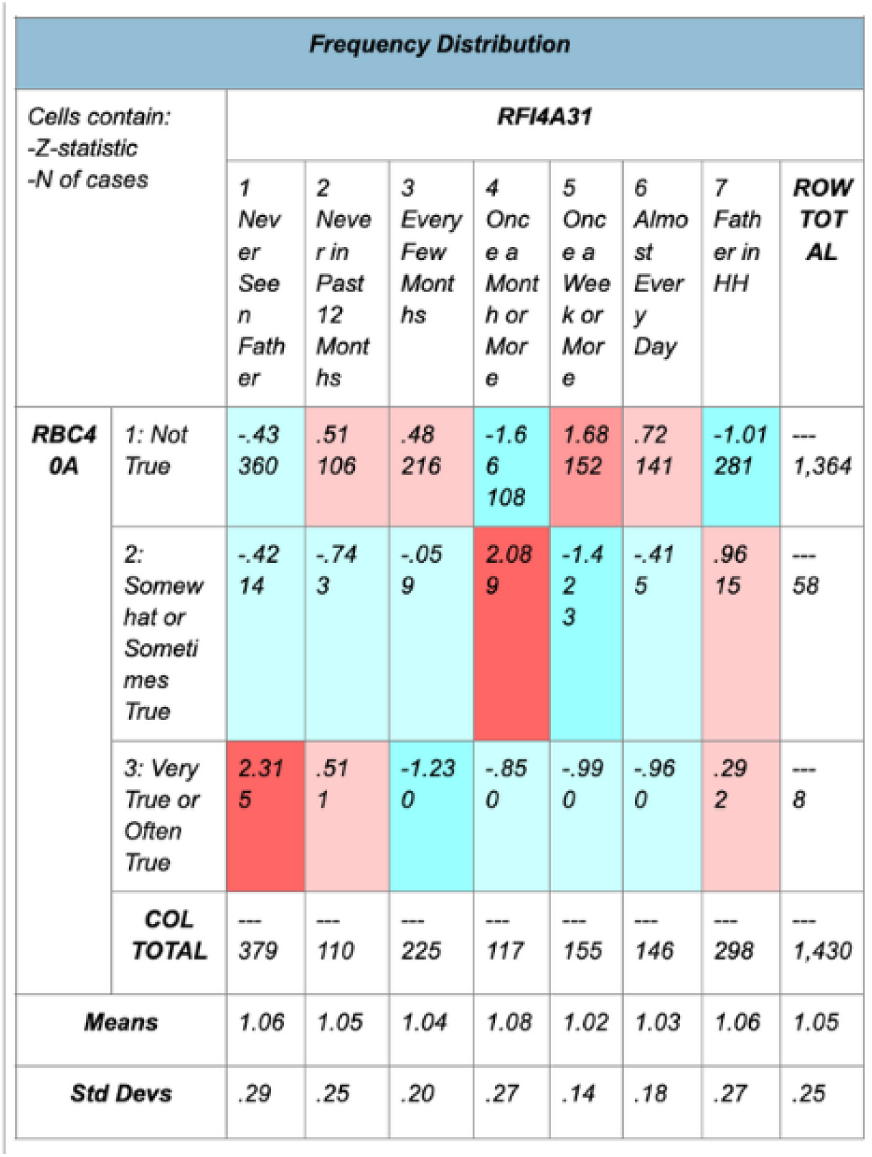
Frequency distribution of child hears voices that aren’t there (RBC40A) and how often the father has seen the focal child in the last 12 months (RFI4A31)

**Figure 15:**
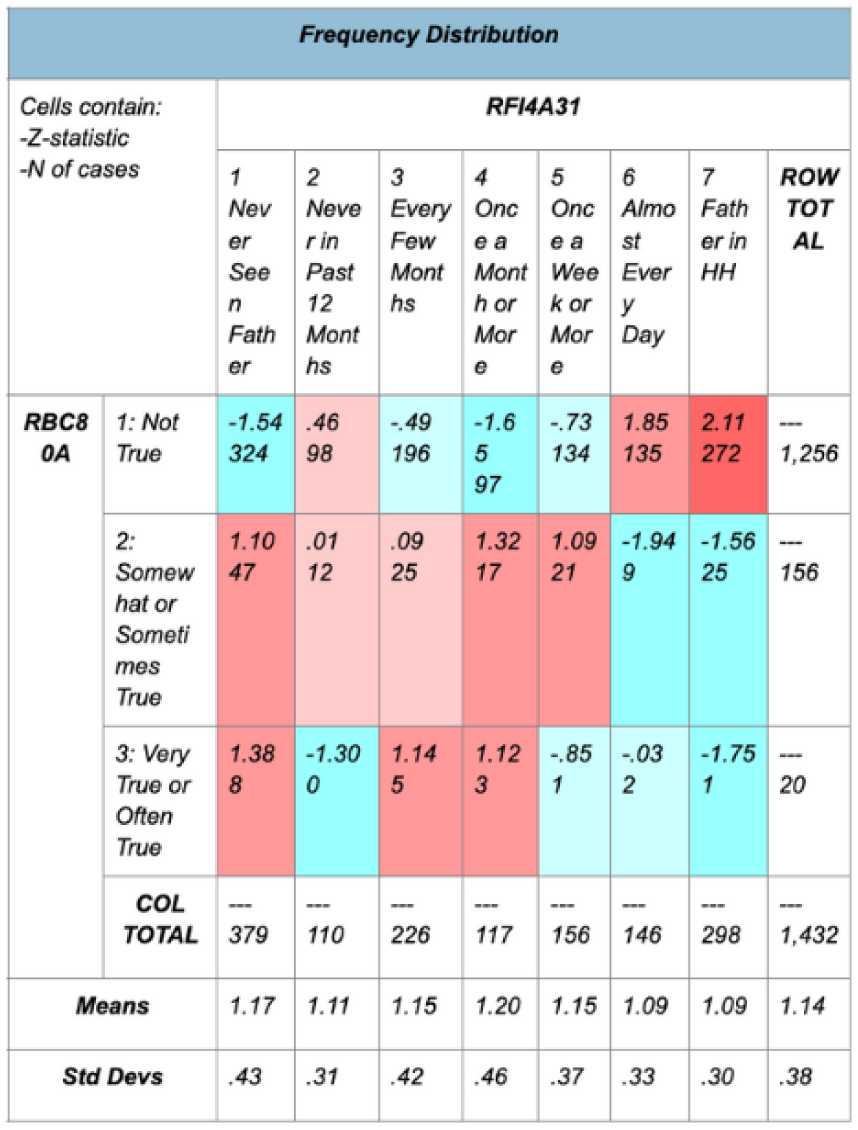
Frequency distribution of child stares blankly (RBC80A) and how often the father has seen the focal child in the last 12 months (RFI4A31)

Figure 1: Children will struggle to concentrate the more violence there is in their neighborhood.

Figure 2: no significant relationship between the child being confused/in a fog and neighborhood violence.

Figure 3: No significant relationship between the child daydreaming or being lost in thought and neighborhood violence.

Figure 4: No significant relationship between a child hearing voices that aren’t there and neighborhood violence.

Figure 5: Children who lived in neighborhood’s with more violence were more likely to be observed staring blankly.

Figure 6: Children were more likely to struggle with concentration the less often they talked to their father in the last 12 months.

Figure 7: Trending but not a significant relationship between how often the father talked to the child and if the child was observed being confused/in a fog. The less often they saw their father, the more like they were to be confused/in a fog.

Figure 8: Trending but not a significant relationship between how often the father talked to the child and how often the child was observed daydreaming or lost in thought. The less often they saw their father the more often they’d be observed lost in thought.

Figure 9: Trending but not a significant relationship between how often the father talked to the child and if the child heard voices that were not there. The less often they saw their father, the more likely they were to hear voices.

Figure 10: Trending but not a significant relationship between how often the father talked to the focal child and if the child was observed staring blankly. The less often they talked to their father, the more likely they were to be staring blankly.

Figure 11: Children were more likely to struggle with concentration the less they saw their father in the last 12 months.

Figure 12: Trending but no significant relationship between how often the father sees the focal child and the child being observed in a fog or confused. The less often they saw their father, the more likely they were to be in a fog.

Figure 13: No significant, but trending relationship between how often the father has seen the child in the last 12 months and if the child was observed daydreaming or lost in thought. The less often they saw their father, the more they would daydream or be lost in thought.

Figure 14: Children who saw their father less often were more likely to hear voices that weren’t there and children who saw their father more often were less likely to hear voices that weren’t there.

Figure 15: Trending but no significant relationship between how often the father has seen the child in the last 12 months and if the child was observed staring blankly. The less often they saw their father, the more likely they were to be staring blankly.

